# Mental distress among Norwegian adults during the Covid-19 pandemic: predictors of initial response and subsequent trajectories

**DOI:** 10.1101/2023.02.06.23285436

**Authors:** Li Lu, Laurie J. Hannigan, Ragnhild E Brandlistuen, Ragnar Nesvåg, Lill Trogstad, Per Magnus, Anna Bára Unnarsdóttir, Unnur A. Valdimarsdóttir, Ole A Andreassen, Helga Ask

## Abstract

**Background:** Understanding factors associated with mental distress during a pandemic is imperative for planning interventions to reduce the negative mental health impact of future crises. Our aim was to identify factors associated with change in levels of mental distress in the Norwegian adult population at the onset of the Covid-19 pandemic, relative to pre-pandemic levels, and with longitudinal changes in mental distress until vaccination against Covid-19 became widespread in Norway (the first 1.5 years of the pandemic).

**Methods:** The Norwegian Mother, Father and Child Cohort Study (MoBa) is a prospective longitudinal study with baseline recruitment from 1999-2009. Baseline characteristics and eight waves of data collection during the pandemic (between March 2020 and September 2021) were used for this analysis. Mental distress was measured with the 5-item version of Hopkins Symptoms Checklist (HSCL-5). A piecewise latent growth model was fitted to identify initial change in mental distress (March-early April 2020, adjusting for pre-pandemic mental distress measured during prior years of data collection) and longitudinal changes across the pandemic in three distinct periods.

**Findings:** Our sample consisted of 105 972 adult participants (59.6% females). Mental distress levels peaked at the beginning of the pandemic. Several factors were associated with initial increases in distress: chronic medical conditions, living alone, history of psychiatric disorders, relatively lower educational background, female sex, younger age, and obesity. Several of these factors were also associated with long-term change. Being quarantined or having to isolate was associated with the likelihood of increasing distress during the pandemic. We observed a reduction in distress associated with Covid-19 vaccination status, while being infected with SARS-CoV-2 was associated with increasing distress late in the pandemic.

**Interpretation:** Pre-pandemic vulnerability factors – like having a chronic disease – as well as Covid-19-related factors – like being quarantined or infected by SARS-CoV-2 – were associated with increased mental distress during the pandemic. This knowledge is important for planning of interventions to support vulnerable individuals during pandemics and other health crises.

**Funding:** The Norwegian Ministry of Health, and Care Services and the Ministry of Education and Research. NordForsk, The Research Council of Norway, The South-Eastern Norway Regional Health Authority.

## Introduction

Being infected with severe acute respiratory syndrome coronavirus 2 (SARS-CoV-2) has been associated with worsened mental health, particularly for the most severely infected (i.e., being hospitalized or bedridden for several days) ^1–3^. Worsened mental health in the population during the pandemic, regardless of infection status, has also been widely reported; including increases in symptoms of anxiety ^4–8^, depression ^4,6–8^, elevated psychological distress ^3,9–11^, sleep problems ^5,6^, and loneliness ^11,12^. Social and environmental changes necessitated by the pandemic and often imposed by governments might have played a direct or indirect role in driving these changes ^13^. However, with much of the research into the relative importance of SARS-CoV-2 infection and public health measures on mental health during the pandemic cross-sectional in design and based on convenience samples, data from population-based, prospective cohorts are urgently needed.

Identifying factors associated with vulnerability and resilience trajectories of mental distress during the coronavirus (Covid-19) pandemic are key for planning targeted interventions to reduce the negative mental impact of future pandemics and for preventing future global health crises. However, since most existing studies lack data on pre-pandemic mental health, there is no way to ascertain how specific the vulnerability or resilience-associated factors they identify are to mental health changes associated with the pandemic; nor to rule out reverse causation. One study showed that female gender, young age, lower income and educational attainment, living alone and having pre-existing mental health conditions were risk factors for anxiety and depression at the start of lockdown, differences were still evident 20 weeks later ^8^. Yet, the study lacked comparable pre-pandemic data, meaning that it was possible that these factors were generally associated with mental health changes across time, and not specifically predictive of changes in the context of the pandemic. The use of large population-based longitudinal data, with long follow-up periods, and rich individual-level information on pre-pandemic mental health can help address this problem.

Using the prospectively and contemporaneously collected data from over 100 000 participants of the Norwegian Mother, Father and Child Cohort Study (MoBa), we first aimed to estimate initial and longitudinal changes in mental distress in the Norwegian adult population during the Covid-19 pandemic, from onset to majority vaccination. At different points during the pandemic, the Norwegian government issued several orders and restrictions, such as closure of schools, remote work, stricter border controls, etc ^14^. Norway went back to normal everyday life in September 2021, 18 months after employing restrictions^14^ and shortly after reaching the milestone of seeing the majority of its population fully vaccinated against the disease. Next, we aimed to identify factors that put individuals at increased risk of worse trajectories of mental health during the pandemic. Crucially, with adjustment for pre-pandemic levels of mental distress based on the same measures of symptomatology, we could estimate the role of other predictors on mental distress trajectories during the pandemic while controlling for individuals’ average levels of pre-pandemic mental distress.

Based on the existing literature^8,15^, we hypothesized that level of mental distress would be elevated at the initial stage of the pandemic, with recovery over time, and pandemic exposures such as income loss, SARS-CoV-2 infections, or quarantine experience would be associated with increases in the level of mental distress experienced during the pandemic.

## Methods

MoBa is an ongoing, population-based pregnancy cohort study conducted by the Norwegian Institute of Public Health ^16,17^. Participants were recruited from all over Norway from 1999-2009. The women consented to participation in 41% of the pregnancies. The cohort now includes 114 500 children, 95 200 mothers and 75 200 fathers. The establishment of MoBa was based on a license from the Norwegian Data Protection Agency and approval from The Regional Committees for Medical and Health Research Ethics. The MoBa cohort is now based on regulations related to the Norwegian Health Registry Act. The current study was approved by The Regional Committees for Medical and Health Research Ethics (14140). All survey participants provided informed consent. In the current study we use pre-pandemic data from version 12 of the quality-assured data files released for research in January 2019.

From March 2020, web-based questionnaires were sent to all adult MoBa participants every second week to collect Covid-19-related information. Participants reported their mental distress in eight waves of data collection (shown alongside the average daily number of new cases of SARS-CoV-2 and level of national mitigating strategies to alleviate the Covid-19 pandemic in Norway in Figure 1). Wave 1 to 8 were respectively responded to from March 31-April 14, 2020, April 14-29, 2020, April 29-May 12, 2020, August 19-September 1, 2020, December 8-21, 2020, February 2-17, 2021, April 28-May 11, 2021 and September 16-29, 2021. Details of each data collection (wave) during the pandemic included in this study are described in the Supplement. We included participants with data from at least three of the eight waves of Covid-19 data collections (N = 105 972).

**Figure 1:**
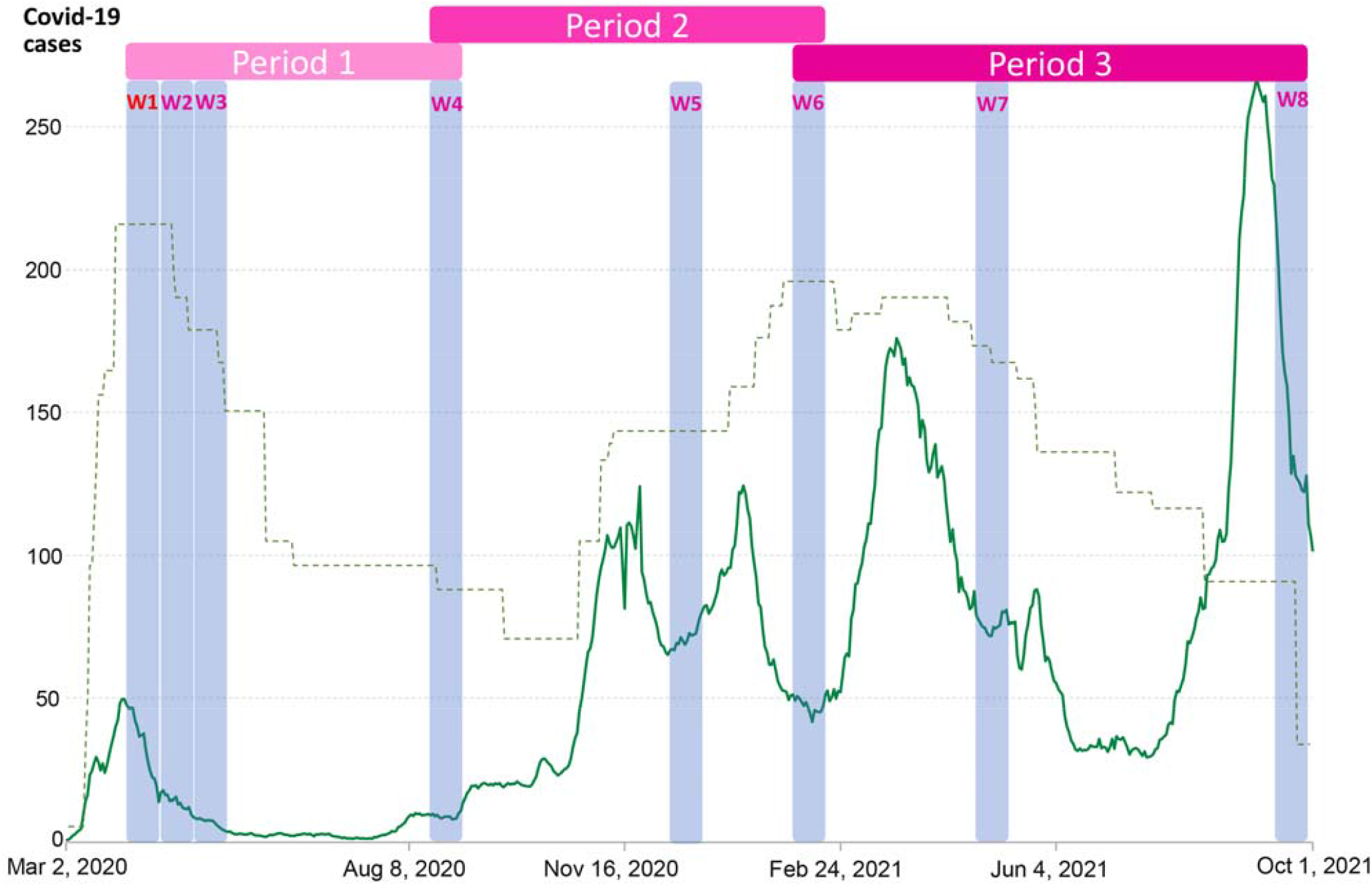
Timing of MoBa Covid-19 data collections (W1-W8) including Hopkins Symptom Checklist-5. **Date of each *wave* (data collection)**: *Wave* 1, March 31-April 14, 2020; *Wave* 2, April 14-29, 2020; *Wave* 3, April 29-May 12, 2020; *Wave* 4, August 19-September 1, 2020; *Wave* 5, December 8-21, 2020; *Wave* 6, February 2-17, 2021; *Wave* 7, April 28-May 11, 2021; *Wave* 8, September 16-29, 2021. **Notes**: The green line shows daily new confirmed Covid-19 cases in Norway per million people. The dotted line indicates Covid-19 Stringency Index – a composite measure based on nine response indicators including school closures, workplace closures and travel bans (unscaled, but show the relative stringency across time periods). Source: Oxford Covid-19 Government Response Tracker, Blavatnik School of Government, University of Oxford. OurWorldInData.org/coronavirus. Periods 1/2/3 are defined according to the general trend in restrictions and in order that each period contains at least 3 waves of data collection for modelling of linear slopes – see the Supplementary Material for further details

## Measures

### Outcomes

*Mental distress* was measured using the 5-item version of the Hopkins Symptoms Checklist (HSCL-5), where participants report their experience of two symptoms of anxiety and three symptoms of depression during the past two weeks ^18^. Each item has 4 response options, ranging from “not at all” (1) to “extremely” (4). HSCL-5 has been widely used and is validated with good psychometric properties in the Norwegian population (Cronbach’s α: 0.87) and in other countries ^19,20^. We excluded records with two or more missing items.

### Covariates

We used the mean of two previous HSCL-5 measures as an indicator of *average levels* of *mental distress across pre-pandemic years*. These included responses on the first available measure, collected between October 1999 and July 2009 (in the mother’s 15^th^ week of gestation) and on the most recent measure before the pandemic (collected when the children were 8 years for the mothers and in year 2015 for the fathers). If data for one measure point was missing, we used the score on the other as indicator of pre-pandemic mental distress.

*Baseline sociodemographic characteristics* included sex, age, education level, and living condition in *Wave* 1, i.e., living alone. Health factors included current Body Mass Index (BMI), which was assessed during the pandemic; history of psychiatric disorders; and chronic medical conditions, that were reported in *Wave* 1-3. *Covid-19-related variables* included income loss, determined by the question included in data collections during the two first time periods; SARS-CoV-2 infection, measured in all waves; and being in quarantine. Information concerning vaccination (SARS-CoV-2) status was available in the surveys *Wave* 6-8. Details of the construction of each variable are reported in the Supplement.

## Statistical analyses

Piecewise latent growth modelling based on data from the eight waves during the pandemic was performed to allow for a nonlinear pattern across time. As a result, our model has one intercept and three slopes, one for each period (see Figure 1 and model illustration in Figure S1). Individual characteristics and health factors were included as predictors of variation in the intercept and all slopes in a single model, equivalent to a multiple regression. In addition, the period-specific Covid-19-related variables were included as predictors for relevant slopes. We ran our model both with and without adjusting for pre-pandemic mental distress, to investigate which predictors were general rather than specific to mental health changes in a pandemic context. All data analyses were performed using *lavaan* version 0.6-9 ^21^ in R version 4.0.0 ^22^ via RStudio ^23^. Full Information Maximum Likelihood estimation was used to handle the missing values ^24^. We used False discovery rate (FDR) to correct for multiple testing.

### Code and data availability

All analytic code is openly available online at https://github.com/psychgen/covid19-adult-mental-distress-trajectories. The consent given by the participants does not open for storage of data on an individual level in repositories or journals. Researchers who want access to datasets for replication should apply through helsedata.no.

## Results

Descriptive statistics are presented in Table 1. The mean mental distress level was highest at the initial stage of the pandemic (mean=7.08, 95% CI=7.06-7.10), and higher than pre-pandemic mental distress (6.09, 95% CI=6.08-6.11). There was an overall pattern of decreasing symptoms of mental distress during the first period of the pandemic, increasing symptoms during the second period, and decreasing symptoms in the final period (Figure S2). Inter-individual variability in mental distress trajectories was substantial, with significant variance terms for each of the growth factors. The model fit was relatively good (i.e., with a root mean square error of approximation (RMSEA) <.06 and comparative fit index (CFI) >.95) ^25^.

**Table 1.** Descriptive characteristics for included sample (N=105 972)

| Variables |  | n | %<sup>a</sup> | Mean (se) |
| --- | --- | --- | --- | --- |
| **Pre-pandemic mental distress** |  | 101 982 | 96,2 | 6,09 (0.01) |
| **Mental distress in** | Wave 1 | 99 851 | 94,2 | 7,08 (0.01) |
|  | Wave 2 | 99 640 | 94,0 | 6,92 (0.01) |
|  | Wave 3 | 97 189 | 91,7 | 6,78 (0.01) |
|  | Wave 4 | 60 736 | 57,3 | 6,34 (0.01) |
|  | Wave 5 | 66 774 | 63,0 | 6,97 (0.01) |
|  | Wave 6 | 82 374 | 77,7 | 6,73 (0.01) |
|  | Wave 7 | 75 670 | 71,4 | 6,59 (0.01) |
|  | Wave 8 | 72 447 | 68,4 | 6,26 (0.01) |
| **Sex:** | Male | 42 800 | 40,4 |  |
|  | Female | 63 172 | 59,6 |  |
| **Age group (years)** | 25-34 | 829 | 0,8 |  |
|  | 35-44 | 36 433 | 34,4 |  |
|  | 45-54 | 61 653 | 58,2 |  |
|  | 55-64 | 6594 | 6,2 |  |
|  | 65 or more | 314 | 0,3 |  |
|  | Missing | 149 | 0,1 |  |
| **Education** | Master or PhD | 30 610 | 28,9 |  |
|  | Bachelor's/university degree | 37 123 | 35,0 |  |
|  | Upper secondary, vocational or other | 30 846 | 29,1 |  |
|  | Compulsory | 2230 | 2,1 |  |
|  | Missing | 5163 | 4,9 |  |
| **BMI** | <25 (Underweight or normal) | 33 978 | 32,1 |  |
|  | 25-30 (Overweight) | 29 358 | 27,7 |  |
|  | >30 (Obese) | 13 433 | 12,7 |  |
|  | Missing | 29 203 | 27,6 |  |
| **History of psychiatric disorders** | Yes | 16 017 | 15,1 |  |
| **N chronic medical conditions** | 1 | 17 315 | 16,3 |  |
|  | 2 | 2428 | 2,3 |  |
|  | >2 | 447 | 0,4 |  |
| **Living alone during the pandemic** | No | 98 672 | 93,1 |  |
|  | Yes | 1114 | 1,1 |  |
|  | Missing | 6186 | 5,8 |  |
| **PANDEMIC-SPECIFIC MEASURES DURING THE FIRST PERIOD** |  |  |  |  |
| **Income loss** | No | 83 394 | 78,7 |  |
|  | Yes, some loss | 14 885 | 14,0 |  |
|  | Yes, significant loss | 6445 | 6,1 |  |
|  | Missing | 1248 | 1,2 |  |
| **SARS-CoV-2 infection** |  | 379 | 0,4 |  |
| **Being quarantined/isolated** |  | 19 560 | 18,5 |  |
| **PANDEMIC-SPECIFIC MEASURES DURING THE SECOND PERIOD** |  |  |  |  |
| **Income loss** | No | 82 077 | 77,5 |  |
|  | Yes, some loss | 16 268 | 15,4 |  |
|  | Yes, significant loss | 7346 | 6,9 |  |
|  | Missing | 281 | 0,3 |  |
| **SARS-CoV-2 infection** |  | 1203 | 1,1 |  |
| **Being quarantined/isolated** |  | 27 620 | 26,1 |  |
| **Vaccinated (SARS-CoV-2)** |  | 2182 | 2,1 |  |
| **PANDEMIC-SPECIFIC MEASURES DURING THE THIRD PERIOD** |  |  |  |  |
| **SARS-CoV-2 infection** |  | 2405 | 2,3 |  |
| **Being quarantined/isolated** |  | 8463 | 8,0 |  |
| **Vaccinated (SARS-CoV-2)** |  | 9323 | 8,8 |  |

BMI: Body Mass Index. N chronic medical conditions: Number of chronic medical conditions.
<sup>a</sup> Values in % column indicate rate of non-missingness in the analytic sample for the first 9 rows, and the breakdown of the sample across groups thereafter.

## Predictors of change in mental distress levels at the onset of the pandemic

### Baseline sociodemographic and health characteristics

The associations of all covariates with initial distress level and the change across the three periods before and after adjusting for pre-pandemic level of distress are shown in Table S1 and Table S2, respectively. Chronic medical conditions (see Table S2 for parameter estimates), living alone (β = 0.28 [SE = 0.03]), female sex (β = 0.11 [0.01]), history of psychiatry disorders (β = 0.17 [0.01]), relative educational background (Table S2) and obesity (β = 0.03 [0.01]) were associated with initial increases in mental distress (i.e., after adjustment for pre-pandemic mental distress). Additionally, being 35-44 years (relative to 45-54), was associated with a slight initial increase (β = 0.04 [0.01]).

Figure 2 shows the differences in standardized association estimates between covariates and initial mental distress before and after adjusting for pre-pandemic mental distress. Virtually all effects were attenuated after adjustment for pre-pandemic mental distress. Effect sizes for history of mental disorders and younger age more than halved after adjustment, rendering the association between young age and initial changes in distress at the onset of the pandemic non-significant.

**Figure 2:**
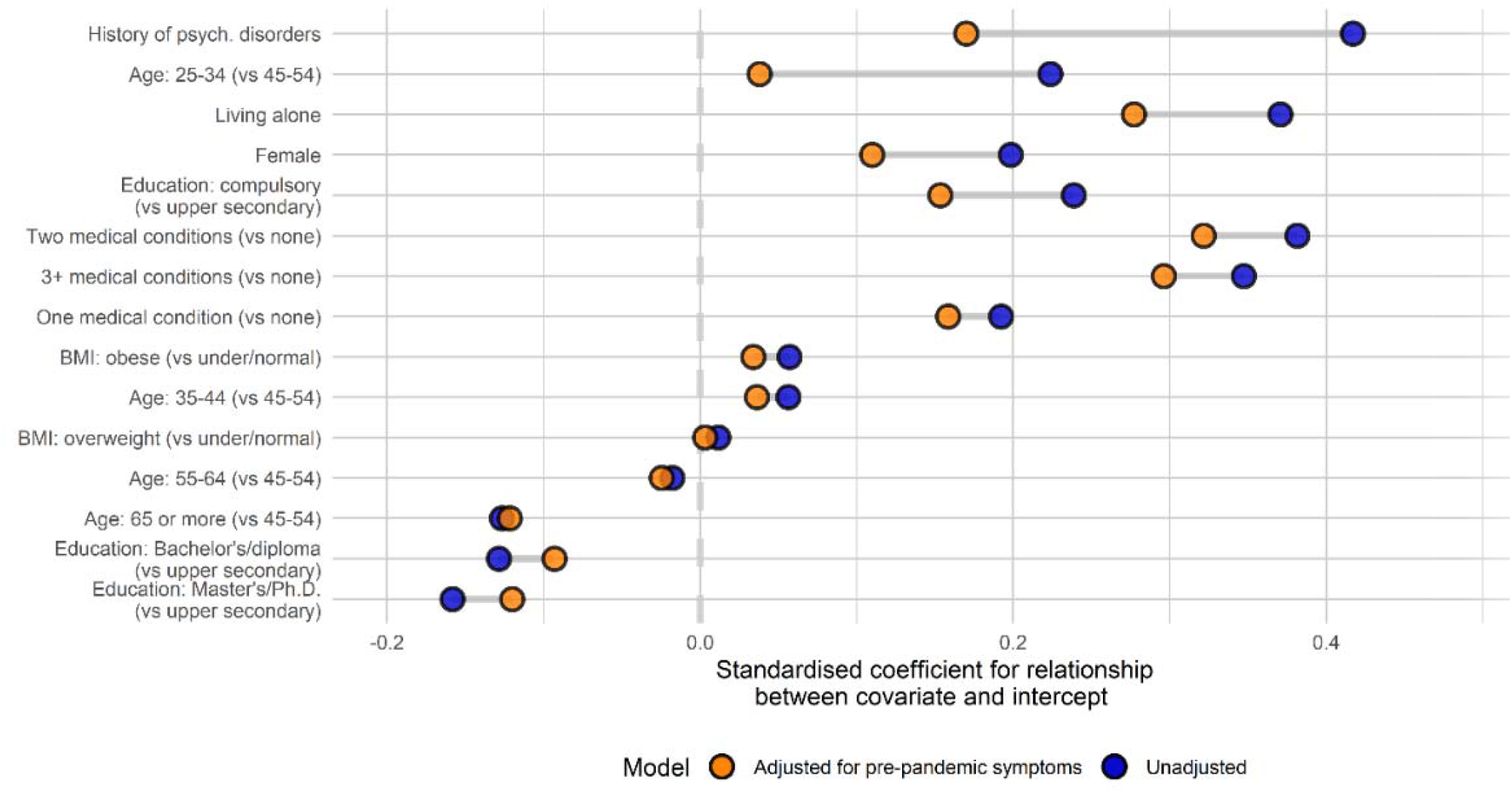
Associations between covariates and initial mental distress before and after adjusting for pre-pandemic mental distress. Note: Covariates are ordered by change in coefficient between the two models. History of psych. Disorders: History of psychiatric disorders; BMI: Body Mass Index.

## Predictors of change in mental distress over the course of the pandemic

### Baseline sociodemographic and health characteristics

Figure 3 shows model-predicted mean HSCL scores stratified according to each baseline characteristic. Sex was the only factor influencing change across all three time periods, with females showing more rapid change in symptoms (β_1_ = −0.08 [0.01]; β_2_ = 0.14 [0.02]; β_3_ = −0.07 [0.01]). Change in symptoms during the first period was also associated with individuals’ educational background, with those with the most education more likely to experience increases in symptoms (β_1_ = 0.13 [0.02]) and those with the least education more likely to experience decreases (β_1_ = −0.10 [0.04]). In addition, those with the highest BMI (β_1_ = 0.06 [0.02]), living alone (β_1_ = 0.14 [0.06]), and with a history of psychiatric disorders (β_1_ = 0.06 [0.02]) were also more likely to experience symptom increases during this period, while those with more chronic medical conditions were more likely to see reductions (having seen, on average, much greater increases at the onset of the pandemic; Table S2). During the second period, the likelihood of experiencing reducing symptoms was associated with older age (particularly being older than 65; β_2_ = −0.28 [0.12]) and living alone (β_2_ = −0.21 [0.07]), reflecting a regression to the mean after prior increases in these groups. The likelihood of symptom increases continued to rise slightly for 34-44 year-olds (relative to those in the age band above; β_2_ = 0.04 [0.01]). The likelihood of symptom decreases during third period was associated with obesity (β_3_ = −0.09 [0.02]), having chronic medical conditions (Table S2), again likely reflecting a reversion to the mean after earlier increases. Younger age continued to be associated with the likelihood of symptoms increasing – most notably among the youngest group (25-34 years; β_3_ = 0.16 [0.07]).

**Figure 3:**
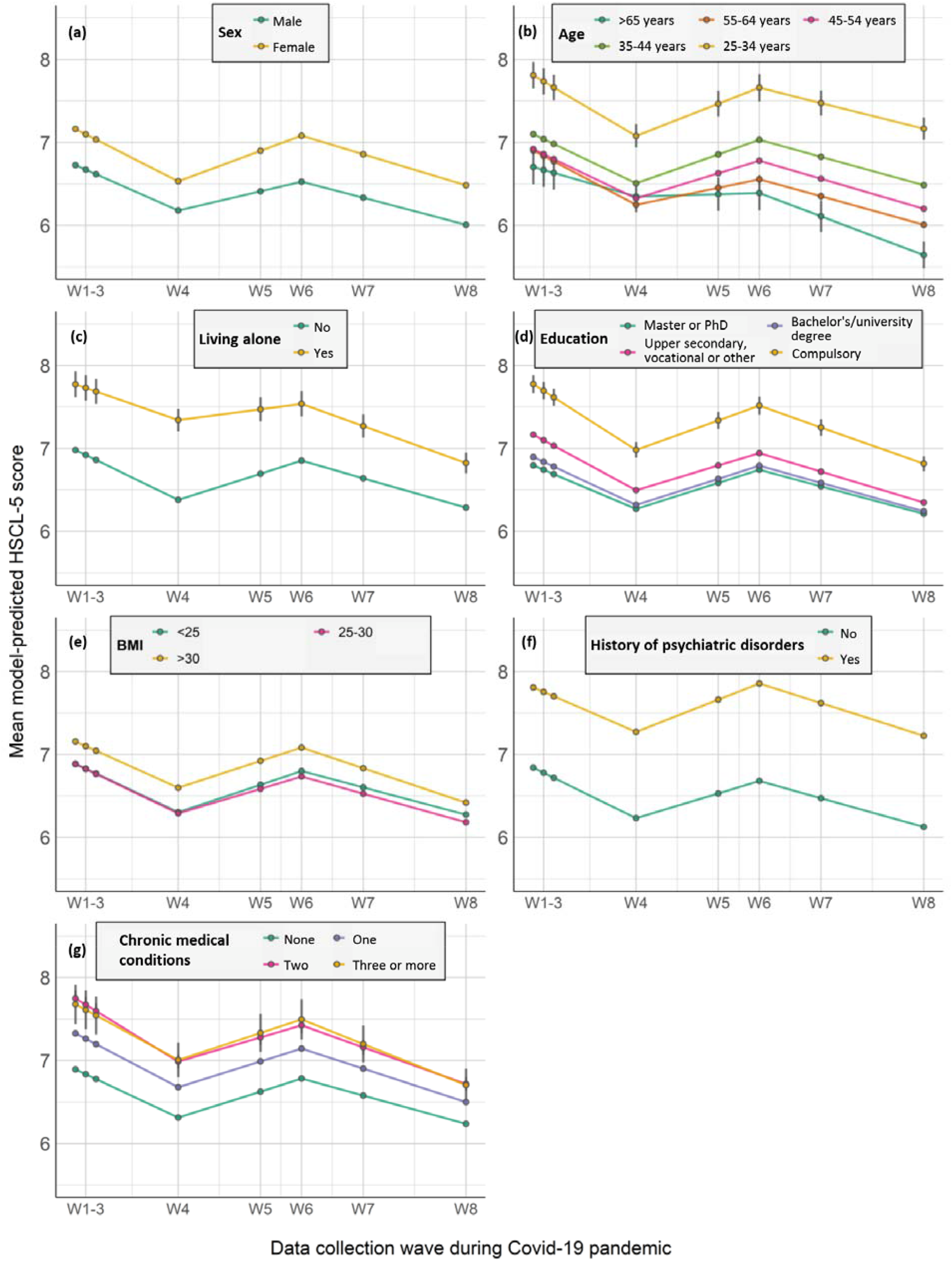
Model-predicted growth trajectories of estimated average mental distress score across waves of data collection (W1-W8) by participant characteristics. Notes: HSCL-5= 5-item version of the Hopkins Symptoms Checklist. Error bar represents 95% confidence intervals; Medical comorbidities= Chronic medical conditions.

### Covid-19-related factors

For the Covid-19-related factors (Figure 4), the likelihood of experiencing increases in mental distress in the first period was associated with significant income loss due to the pandemic in this period (β_1_ = 0.12 [0.02]), as well as being quarantined or having to isolate during this period (β_1_ = 0.08 [0.01]). During the second period, being vaccinated was associated with likelihood of symptom reduction (β_2_ = −0.18 [0.03]) while symptom increases were again more likely among individuals quarantined during this period (β_2_ = 0.11 [0.01]). In the final period, those who suffered income loss during the prior period were more likely to see symptom reduction (β_3_ = −0.15 [0.02]), while SARS-CoV-2 infection was associated with likelihood of symptom increases (β_2_ = 0.10 [0.04]) for the first time (case numbers had remained very low in Norway during the two prior periods).

**Figure 4:**
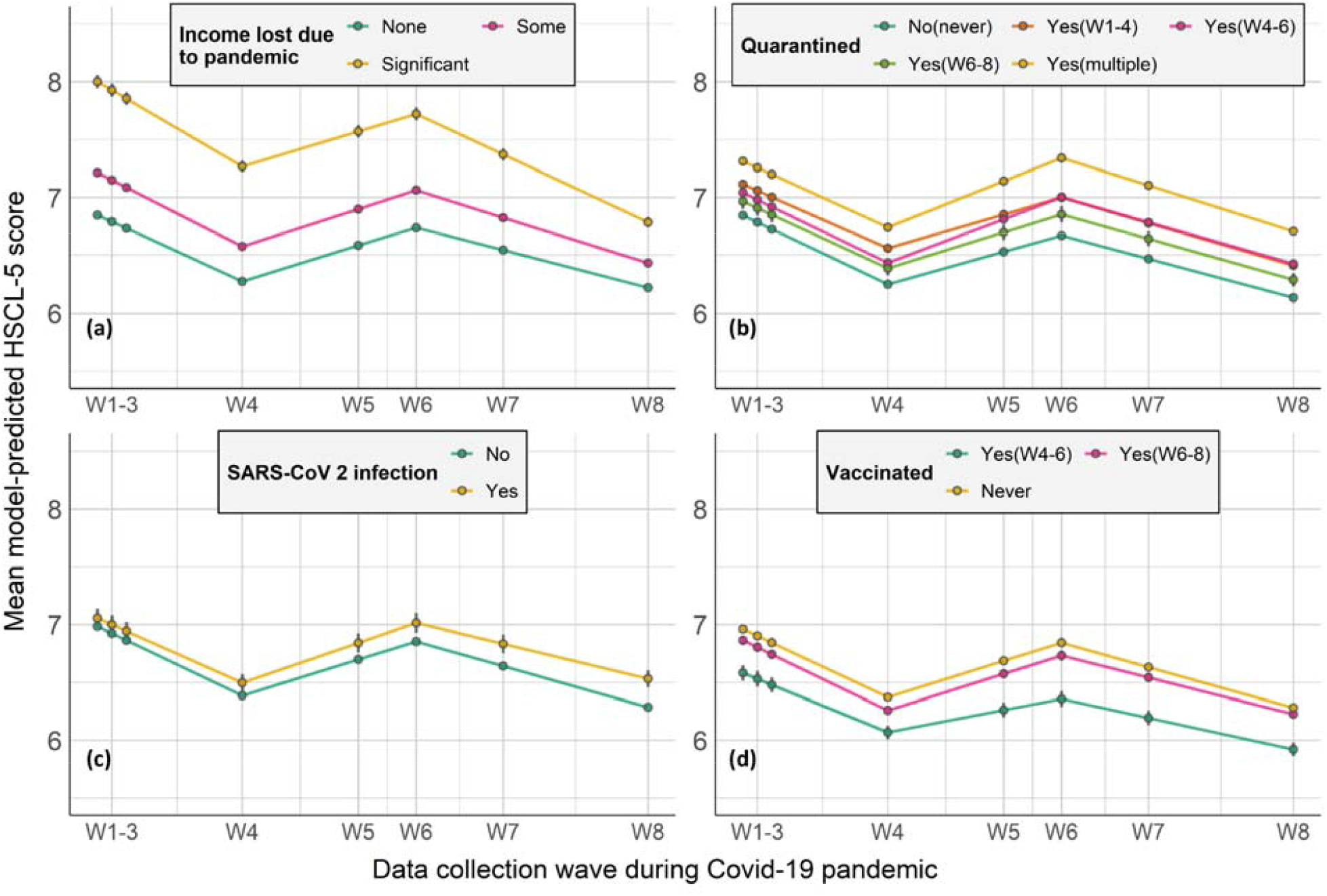
Model-predicted growth trajectories of estimated average mental distress score across waves of data collection (W1-W8) by Covid-19-related factors. Notes: HSCL-5= 5-item version of the Hopkins Symptoms Checklist. Error bar represents 95% confidence intervals.

The effect of adjusting for pre-pandemic mental distress on estimated associations between covariates and change during each period are displayed in Figures S3-S5).

## Discussion

Using prospectively collected data from a large nationwide cohort, we found that several factors were associated with initial increases in mental distress in the beginning of the pandemic. These included having chronic medical conditions or a history of mental disorders, relative lower educational background, female sex, living alone, and obesity, which were also associated with longitudinal change. Being quarantined and losing income due to Covid-19 were associated with likelihood of increasing distress during the first year of the pandemic. We found that having received the Covid-19 vaccination was associated with decreasing symptoms during the second period while SARS-CoV-2 infection was independently associated with increasing symptoms of mental distress only towards the end of the study period, which coincided with the first substantial rise in case numbers in Norway.

To isolate factors not just related to mental health in general, but to changes in mental health during the pandemic, we adjusted our models for pre-pandemic mental distress. A majority of studies regarding mental health and Covid-19 so far have been based on online recruitment and data collected after the start of the pandemic ^7,8,26^, without knowledge of pre-pandemic mental distress. We found the strength of associations with mental distress during the pandemic are reduced after adjustment for pre-pandemic data, particularly for variables such as age, history of psychiatric disorders, living alone, sex, education and chronic medical conditions. Findings from previous studies that did not control for these factors are likely to conflate general and specific effects on mental health and should be interpreted as such. To the extent that such findings are interpreted as being specific to the context of the pandemic, our results show that they are likely to be substantially inflated.

Our findings show that female sex and younger age were associated with higher initial level and more change in mental distress across time. This is in line with the results from a UK study performing growth models on anxiety- and depressive symptoms ^8^. It is documented that women are more likely to suffer from mental health problems than men ^4,9,12,13^, and females had higher increase than males in prevalence of major depressive and anxiety disorders during Covid-19, particularly in younger age groups ^13^. During the pandemic, additional carer and household responsibilities due to school closures or family members becoming unwell are more likely to fall on women ^13,27^. We also found that women demonstrated faster improvements in anxiety and depression symptoms during the first period.

Living alone was associated with both a higher initial level and less rapid recovery during the first period. Factors like living alone and loneliness have repeatedly been linked to mental health ^12^, also before the pandemic ^28^. Interestingly, we observed that being older than 65 was associated with a relative reductions in risk for mental distress at the onset of, and during the pandemic. This was despite older age being a known risk factor for more severe disease from relatively early in the pandemic. Interacting sociodemographic factors, such as relative insulation from economic concerns or living in less densely populated areas and potential increase in social support may explain this somewhat counter-intuitive finding.

Individuals with chronic medical conditions presented with a substantial increase in mental distress at the initial stage of the pandemic, showed reversion towards the mean over time, similar to the UK study ^8^. Obesity was slightly predictive of increases in mental distress at the onset and for the early phase of the pandemic, and reversion toward the mean came later, during the period when vaccination became widespread. Obesity and chronic medical conditions are risk factors for Covid-19 severity ^29^, which was also communicated extensively during the first stages of the pandemic. This, followed by the implementation of greater shielding and infection control measures, as well as adjustment to perceived vulnerability, could explain the early increases and subsequent decreases in mental distress for these factors. Our findings of pre-existing mental illness as a significant predictor of mental problems were expected as it has been widely reported ^3,30,31^, although again we confirm that this is specific to the pandemic context by appropriately controlling for earlier symptoms. It is also worth noting that in contrast to medical comorbidities and obesity, individuals with pre-existing vulnerabilities to psychiatric disorders showed no reversion to the mean at all during our study period, suggesting that individuals with these vulnerabilities should be prioritised for support in analogous future situations.

Covid-19-related characteristics, such as loss of income and being quarantined were associated with slower improvement during the first period. Economic loss is one significant adversity in the context of Covid-19, and could be where Covid-19 related stress partially originates from ^32^. Being quarantined was associated with a greater likelihood of experiencing increases in symptoms during the second period. In addition to the obvious possibility that people were concerned that exposure to infected individuals might lead to them developing the disease, social isolation to mitigate the spread of the novel coronavirus may have led to loneliness, which is associated with increased mental health problems ^33^. During the second period, the likelihood of seeing symptoms of distress decrease was associated with being vaccinated. This protective effect of vaccination has been observed previously ^34^. Decrease in mental distress symptoms during the third period was more rapid among those with significant income loss in the previous period. The greater improvement of those with income loss could be partially explained by the government mitigation strategies including unemployment benefit, i.e., more compensation would be offered under new rules ^14^; and possible easing of economic and employment pressures. Individuals infected with SARS-CoV-2 were more likely to experience increased mental distress during the third period than those not infected. A negative association between SARS-CoV-2 infection and mental health have been observed in several studies ^2,3,13^ while our study extends the literature by robustly controlling for pre-pandemic levels of distress.

Our findings extend current knowledge and provide important implications for future studies by taking the advantage of the longitudinal design and pre-pandemic information available in the MoBa cohort with data between the onset of the Covid-19 pandemic and majority vaccination against Covid-19. However, the results should be interpreted in light of some limitations. First, the current study only included MoBa parent participants, therefore the results cannot necessarily be generalised to the adult population without children. The initial participation rate in MoBa was 41% of invited pregnant women, with previously described overrepresentation of healthier and more highly educated women as compared to the general population^35^. A second limitation concerns our inability to draw any strong causal conclusions about links between individual characteristics and trajectories of mental distress based on observational data.

In conclusion, our results identify several vulnerability factors, including living alone, obesity, history of psychiatry disorders that represent risk factors for both initial and longitudinal increases in mental distress in the context of the Covid-19 pandemic. Significant influences, too, of quarantine status, vaccination, and income loss due to the pandemic show that public health and governmental policies and priorities are likely to be influential at the level of individual mental health among citizens. These findings are important for planning of future public health responses to reduce the mental health impact of Covid-19 and similar future global health crises, as well as to optimize the allocation of health service resources and the design of prevention and intervention efforts.

## Data Availability

The consent given by the participants does not open for storage of data on an individual level in repositories or journals. Researchers who want access to datasets for replication should apply through helsedata.no.

https://helsedata.no/

## Acknowledgement

We thank all the participating families in Norway who take part in the ongoing MoBa cohort study. The authors are grateful for technical support in statistics from Dr. Baeksan Yu.

## Contributors

REB, LT and HA involved in designing the data collections of the MoBa cohort. LL, LJH and HA designed the analytical strategy and all authors helped to interpret the findings. LL, LJH and HA conducted the literature review and drafted the manuscript. All authors revised the manuscript for critical content and approved the final version of the manuscript. LJH and HA had access to and verified all the data and all authors could access the data on request.

