## Supplementary material for "Mental distress among Norwegian adults during the Covid-19 pandemic: predictors of initial response and subsequent trajectories": Online supplementary materials

### Methods

##### Details of each data collection (wave) during the pandemic included in this study

*Wave 1* (responded to from March 31-April 14, 2020) was collected right after the pandemic mitigation measures in Norway started in March 2020, with several restrictions issued including physical distancing, working from home order, closure of schools, nonessential businesses, and many public facilities ^1^. Government support schemes were gradually introduced, and restrictions relieved during *Wave 2* (April 14-29, 2020) and *Wave 3* (April 29-May 12, 2020). The lockdown measures were relaxed in the summer 2020 and before *Wave 4* data collection (August 19-September 1, 2020). New measures were gradually re-introduced throughout December, 2020 ^1^ and during the time of data collection *Wave 5* (December 8-21, 2020). In December 2020, the Covid-19 vaccination program in Norway also started, with large parts of the oldest population vaccinated by *Wave 6* (February 2-17, 2021). In March 11, 2021, around 5% of the population had been vaccinated with the second dose ^2^. During *Wave 7* (April 28-May 11, 2021) around 10% of the population were fully vaccinated and there were 75 daily new confirmed cases of SARS-CoV-2 per million people. At the time of *Wave 8* (September 16-29, 2021) around 67% of the population were fully vaccinated with 100-200 daily new confirmed SARS-CoV-2 cases per million people ^2^.

##### Covariates

*Baseline sociodemographic characteristics* included sex (female/male), age (25-34 years/35-44/45-54/55-64/65 or more), education level (compulsory/upper secondary, vocational or other/bachelor’s, diploma or similar university degree/master’s or PhD), and living condition in *Wave 1*, i.e., living alone (no/yes).

*Health factors* included current Body Mass Index (BMI), which was assessed during the pandemic and calculated by dividing the body weight by squared height (kilograms/meter^2^) and categorised to underweight or normal (<25), overweight (25-30) and obese (>30) ^3^; History of psychiatric disorders (no/yes) was constructed by combining self-reported information from pre-pandemic questionnaires: ‘Do you have or have you had any of the following illnesses or health problems (Depression/anxiety/anorexia and bulimia/manic depressive illness/schizophrenia/other long-term mental illnesses/bipolar disorder/mental disorder)?’, and the Life Time History of Major Depression ^4^; chronic medical conditions (no/one/two/three or more conditions) were reported in Wave1-3: ‘Do you have any of the following diseases (Asthma or other lung disease/Cancer/Heart disease/Diabetes/ Hypertension)?’

*Covid-19-related variables* included income loss, determined by the question ‘Have you lost income as a result of the Covid-19 pandemic (no/yes, some/yes, significant)?’, included in data collections during the two first time periods; SARS-CoV-2 infection (no/yes), measured in all waves by the questions ‘Have you been tested for coronavirus infection during the last 14 days (no/yes)’ and ‘Did the test confirm coronavirus infection (no/yes)?’ and it was defined as with SARS-CoV-2 infection if at least one reply with ‘yes’; Being in quarantine during each time period (yes/no) was defined as with quarantine in at least one data collection in each period. Information concerning vaccination (SARS-CoV-2) status (no/yes) was available in the surveys Wave 6-8.

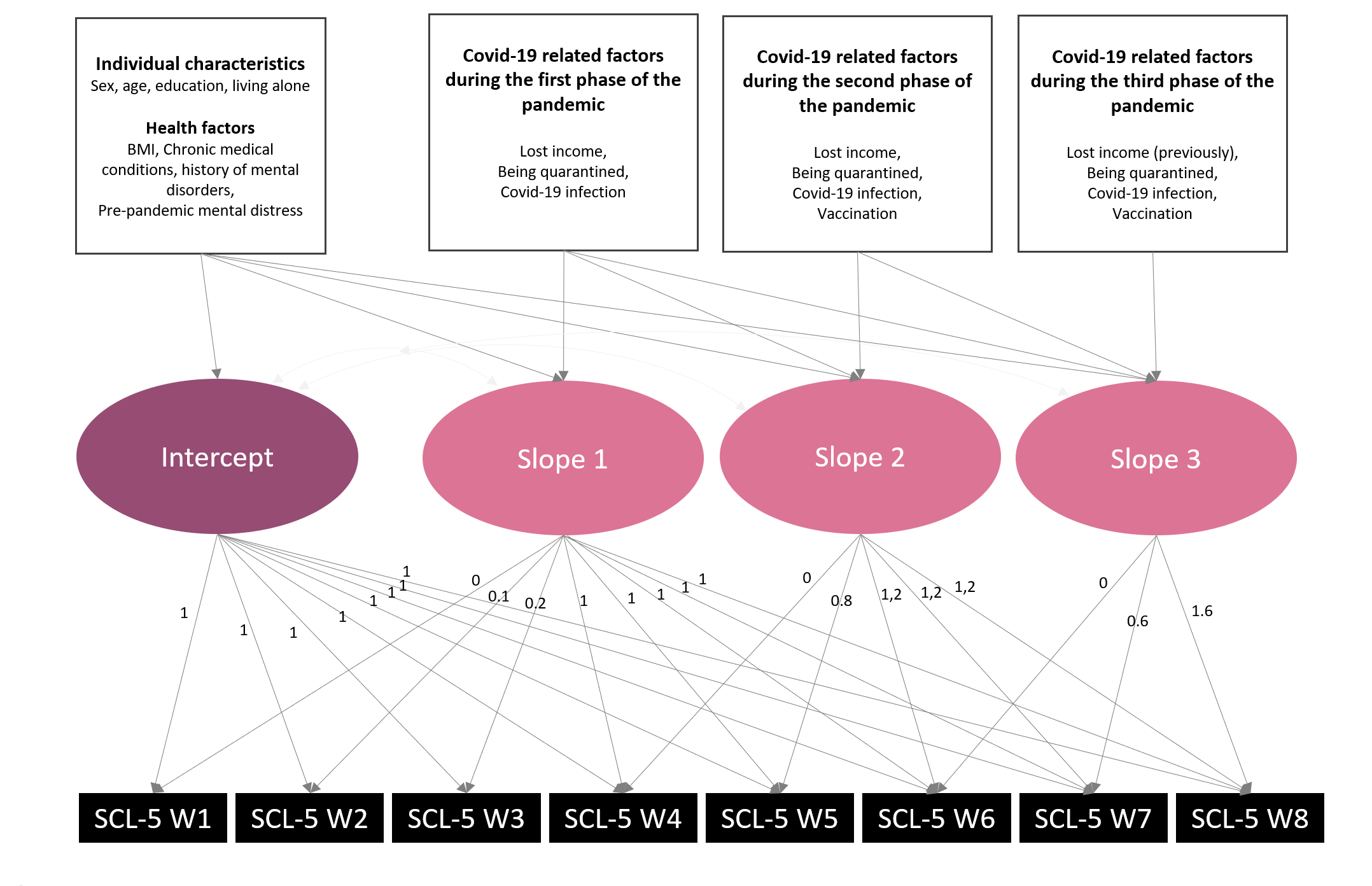

### Figure S1 – Piecewise growth model - illustration

*Note: Squares represents measured variables, circles represent latent factors. SCL-5= Hopkins symptom checklist-5; W1-W8= Data collection wave 1-8;*

Piecewise latent growth modelling requires at least three waves of data for each “piece” (hereafter: “period”), and at least one wave of overlap with the preceding/subsequent period. As such, with eight waves of data available to us overall, we could model slopes for three periods, one of which would necessarily include four waves of data (with three waves included in each of the others). We included four waves in the first period (Waves 1-4), since Wave 1-3 were so close together in time, and since there was a general pattern of releasing of restrictions during this period (see Figure 1). The second and third periods consisted of Wave 4-6 and Wave 6-8, respectively.

### Figure S2 – Model-predicted mean across pandemic waves (W1-W8)

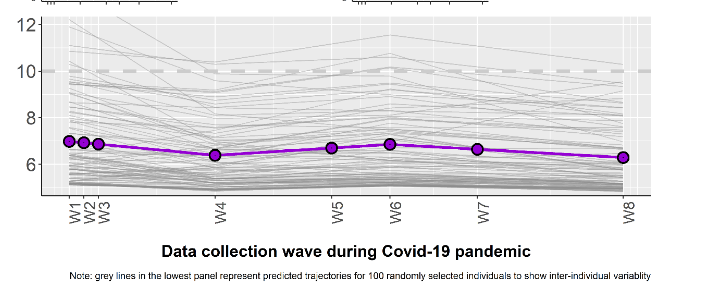

Note: The grey lines represent predicted trajectories for 100 randomly selected individuals to show the inter-individual variability. The dotted line represents a cutoff commonly used to indicate clinical significance on this scale.

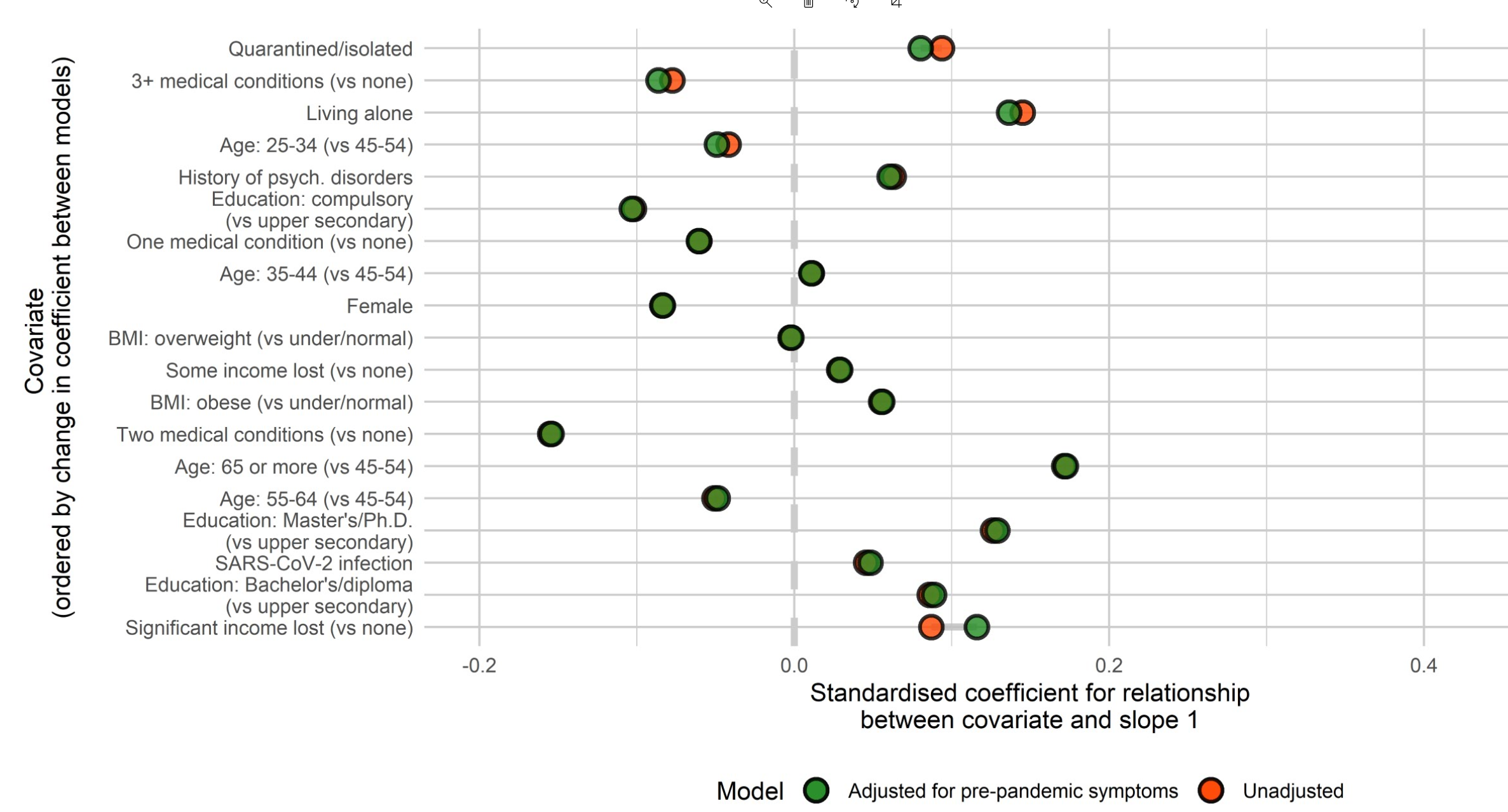

### Figure S3 – Associations between covariates and change during the first period (slope 1) before and after adjusting for pre-pandemic mental distress

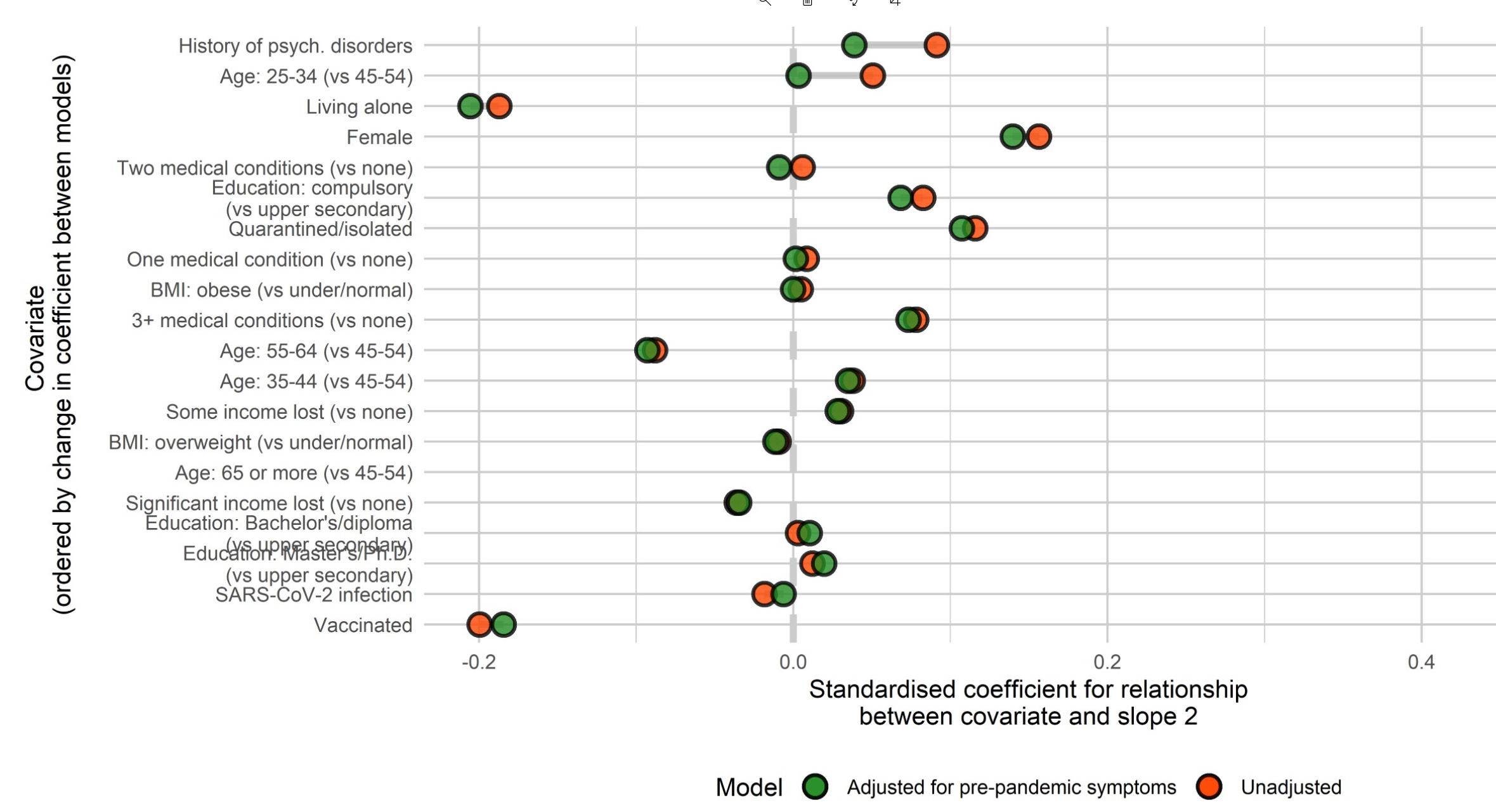

### Figure S4 – Associations between covariates and change during the second period (slope 2) before and after adjusting for pre-pandemic mental distress

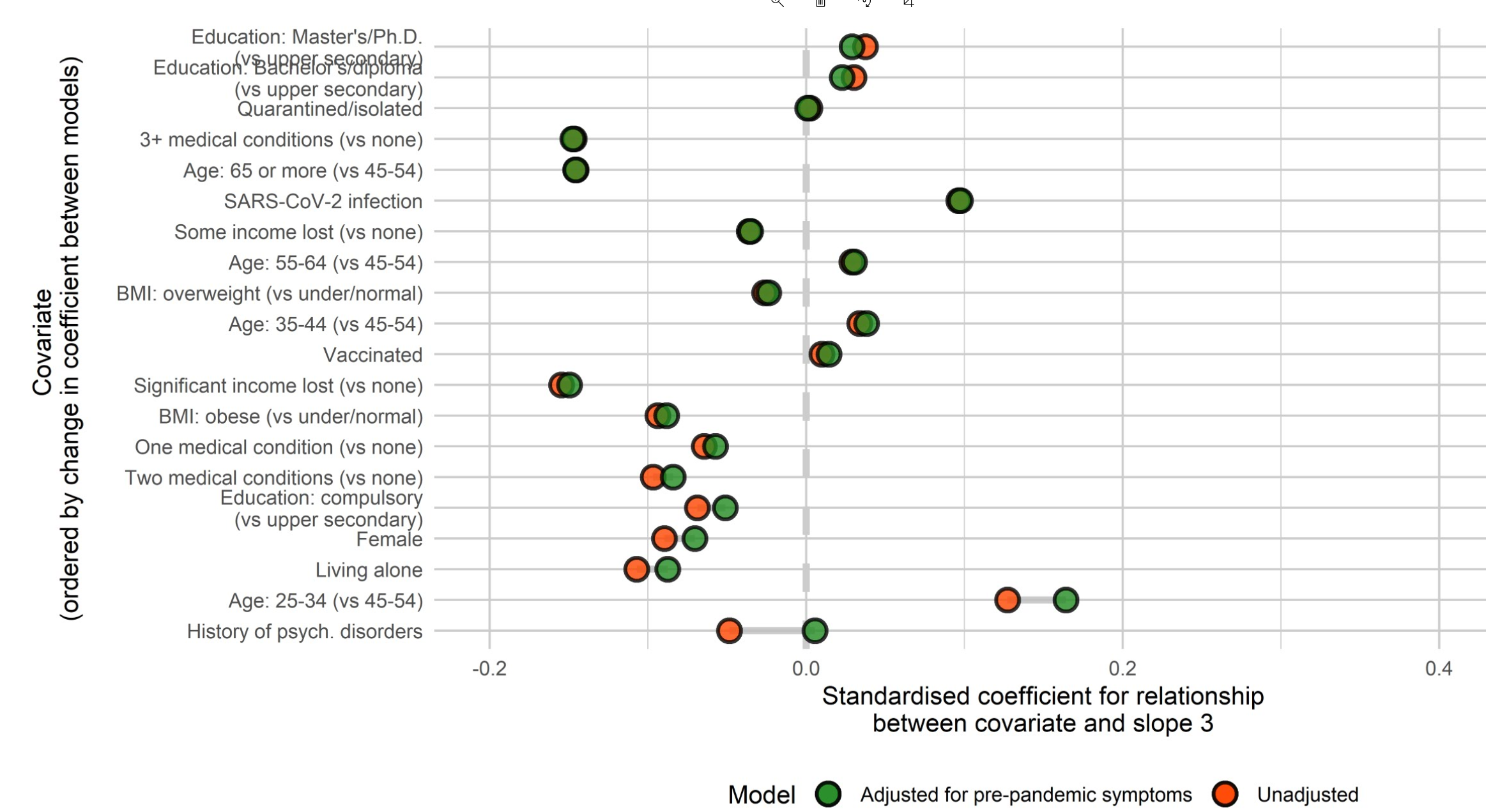

### Figure S5 – Associations between covariates and change during the second period (slope 3) before and after adjusting for pre-pandemic mental distress

### Table S1: Estimated effects of the covariates on the intercepts and slopes from the piecewise growth model (n=105 972) without adjusting for pre-pandemic mental distress

|  | **Predictors of the intecept** | | | |  | **Predictors first slope** | | | |  | **Predictors of second slope** | | | |  | **Predictors of third slope** | | | |
| --- | --- | --- | --- | --- | --- | --- | --- | --- | --- | --- | --- | --- | --- | --- | --- | --- | --- | --- | --- |
| **Covariate** | **Estimate** | **SE** | **p-value** | **FDR-p** |  | **Estimate** | **SE** | **p-value** | **FDR-p** |  | **Estimate** | **SE** | **p-value** | **FDR-p** |  | **Estimate** | **SE** | **p-value** | **FDR-p** |
| **Sex** |  |  |  |  |  |  |  |  |  |  |  |  |  |  |  |  |  |  |  |
| Female (vs male) | 0,198 | 0,007 | 0,000 | 0,000 |  | -0,084 | 0,012 | 0,000 | 0,000 |  | 0,156 | 0,014 | 0,000 | 0,000 |  | -0,090 | 0,013 | 0,000 | 0,000 |
| **Age (years)** |  |  |  |  |  |  |  |  |  |  |  |  |  |  |  |  |  |  |  |
| 25-34 (vs 45-54) | 0,224 | 0,037 | 0,000 | 0,000 |  | -0,042 | 0,064 | 0,515 | 0,586 |  | 0,051 | 0,076 | 0,504 | 0,586 |  | 0,127 | 0,068 | 0,061 | 0,098 |
| 35-44 (vs 45-54) | 0,056 | 0,007 | 0,000 | 0,000 |  | 0,011 | 0,012 | 0,365 | 0,458 |  | 0,038 | 0,014 | 0,009 | 0,018 |  | 0,034 | 0,013 | 0,009 | 0,018 |
| 55-64 (vs 45-54) | -0,018 | 0,014 | 0,190 | 0,246 |  | -0,051 | 0,024 | 0,033 | 0,057 |  | -0,088 | 0,028 | 0,002 | 0,005 |  | 0,029 | 0,025 | 0,255 | 0,326 |
| 65 or more (vs 45-54) | -0,127 | 0,059 | 0,032 | 0,057 |  | 0,171 | 0,103 | 0,097 | 0,141 |  | -0,279 | 0,122 | 0,022 | 0,041 |  | -0,145 | 0,110 | 0,185 | 0,244 |
| **Education** (vs upper secondary/vocational) |  |  |  |  |  |  |  |  |  |  |  |  |  |  |  |  |  |  |  |
| Compulsory | 0,239 | 0,023 | 0,000 | 0,000 |  | -0,102 | 0,040 | 0,011 | 0,021 |  | 0,083 | 0,047 | 0,080 | 0,120 |  | -0,069 | 0,042 | 0,103 | 0,147 |
| Bachelor’s/university degree | -0,129 | 0,008 | 0,000 | 0,000 |  | 0,086 | 0,014 | 0,000 | 0,000 |  | 0,003 | 0,017 | 0,859 | 0,890 |  | 0,030 | 0,015 | 0,046 | 0,076 |
| Master/PhD | -0,158 | 0,009 | 0,000 | 0,000 |  | 0,126 | 0,015 | 0,000 | 0,000 |  | 0,012 | 0,018 | 0,499 | 0,586 |  | 0,037 | 0,016 | 0,019 | 0,036 |
| **BMI (kg/m^2)** (vs <25 (Underweight or normal)) |  |  |  |  |  |  |  |  |  |  |  |  |  |  |  |  |  |  |  |
| 25-30 (Overweight) | 0,012 | 0,009 | 0,174 | 0,233 |  | -0,002 | 0,014 | 0,866 | 0,890 |  | -0,009 | 0,016 | 0,560 | 0,626 |  | -0,026 | 0,014 | 0,068 | 0,104 |
| >30 (Obese) | 0,057 | 0,011 | 0,000 | 0,000 |  | 0,055 | 0,018 | 0,002 | 0,006 |  | 0,005 | 0,021 | 0,822 | 0,869 |  | -0,094 | 0,019 | 0,000 | 0,000 |
| **History of psychiatric disorders** |  |  |  |  |  |  |  |  |  |  |  |  |  |  |  |  |  |  |  |
| Yes vs No | 0,417 | 0,009 | 0,000 | 0,000 |  | 0,063 | 0,016 | 0,000 | 0,000 |  | 0,091 | 0,019 | 0,000 | 0,000 |  | -0,049 | 0,017 | 0,004 | 0,009 |
| **Living alone** |  |  |  |  |  |  |  |  |  |  |  |  |  |  |  |  |  |  |  |
| Yes vs No | 0,371 | 0,031 | 0,000 | 0,000 |  | 0,145 | 0,055 | 0,008 | 0,017 |  | -0,187 | 0,065 | 0,004 | 0,009 |  | -0,107 | 0,059 | 0,067 | 0,104 |
| **Chronic medical conditions** |  |  |  |  |  |  |  |  |  |  |  |  |  |  |  |  |  |  |  |
| 1 condition (vs no) | 0,192 | 0,009 | 0,000 | 0,000 |  | -0,060 | 0,015 | 0,000 | 0,000 |  | 0,008 | 0,018 | 0,644 | 0,701 |  | -0,064 | 0,016 | 0,000 | 0,000 |
| 2 conditions (vs no) | 0,381 | 0,022 | 0,000 | 0,000 |  | -0,155 | 0,038 | 0,000 | 0,000 |  | 0,006 | 0,045 | 0,895 | 0,895 |  | -0,097 | 0,040 | 0,016 | 0,032 |
| > 2 conditions (vs no) | 0,347 | 0,050 | 0,000 | 0,000 |  | -0,078 | 0,087 | 0,371 | 0,458 |  | 0,078 | 0,102 | 0,446 | 0,541 |  | -0,147 | 0,092 | 0,111 | 0,155 |
| **Income lost due to the pandemic** |  |  |  |  |  |  |  |  |  |  |  |  |  |  |  |  |  |  |  |
| yes, some (vs no) |  |  |  |  |  | 0,028 | 0,014 | 0,042 | 0,071 |  | 0,030 | 0,018 | 0,086 | 0,127 |  | -0,036 | 0,017 | 0,029 | 0,052 |
| yes, significant (vs no) |  |  |  |  |  | 0,087 | 0,021 | 0,000 | 0,000 |  | -0,036 | 0,025 | 0,155 | 0,213 |  | -0,155 | 0,023 | 0,000 | 0,000 |
| **Quarantined/isolated** |  |  |  |  |  |  |  |  |  |  |  |  |  |  |  |  |  |  |  |
| Yes (vs no) |  |  |  |  |  | 0,094 | 0,010 | 0,000 | 0,000 |  | 0,116 | 0,010 | 0,000 | 0,000 |  | 0,002 | 0,017 | 0,894 | 0,895 |
| **SARS-CoV-2-infection** |  |  |  |  |  |  |  |  |  |  |  |  |  |  |  |  |  |  |  |
| Yes (vs no) |  |  |  |  |  | 0,046 | 0,070 | 0,514 | 0,586 |  | -0,018 | 0,049 | 0,708 | 0,760 |  | 0,096 | 0,036 | 0,007 | 0,016 |
| **Vaccination (SARS-CoV-2) status** |  |  |  |  |  |  |  |  |  |  |  |  |  |  |  |  |  |  |  |
| Yes (vs no) |  |  |  |  |  |  |  |  |  |  | -0,200 | 0,030 | 0,000 | 0,000 |  | 0,010 | 0,017 | 0,567 | 0,626 |

Notes: Model was estimated with full Information Maximum Likelihood method, BMI: Body Mass Index. SE: Standard error; FDR p: False Discovery Rate corrected p-value.

Table S2: Estimated effects of the covariates on the intercepts and slopes from the piecewise growth model (n=105 972) after adjusting for pre-pandemic mental distress

|  | **Predictors of the intecept** | | | |  | **Predictors first slope** | | | |  | **Predictors of second slope** | | | |  | **Predictors of third slope** | | | |
| --- | --- | --- | --- | --- | --- | --- | --- | --- | --- | --- | --- | --- | --- | --- | --- | --- | --- | --- | --- |
| **Covariate** | **Estimate** | **SE** | **p-value** | **FDR-p** |  | **Estimate** | **SE** | **p-value** | **FDR-p** |  | **Estimate** | **SE** | **p-value** | **FDR-p** |  | **Estimate** | **SE** | **p-value** | **FDR-p** |
| **Sex** |  |  |  |  |  |  |  |  |  |  |  |  |  |  |  |  |  |  |  |
| Female (vs male) | 0,110 | 0,007 | 0,000 | 0,000 |  | -0,084 | 0,012 | 0,000 | 0,000 |  | 0,140 | 0,015 | 0,000 | 0,000 |  | -0,070 | 0,013 | 0,000 | 0,000 |
| **Age (years)** |  |  |  |  |  |  |  |  |  |  |  |  |  |  |  |  |  |  |  |
| 25-34 (vs 45-54) | 0,038 | 0,035 | 0,279 | 0,356 |  | -0,049 | 0,064 | 0,444 | 0,533 |  | 0,003 | 0,076 | 0,965 | 0,978 |  | 0,164 | 0,068 | 0,016 | 0,033 |
| 35-44 (vs 45-54) | 0,036 | 0,007 | 0,000 | 0,000 |  | 0,011 | 0,012 | 0,377 | 0,466 |  | 0,035 | 0,014 | 0,016 | 0,033 |  | 0,038 | 0,013 | 0,003 | 0,007 |
| 55-64 (vs 45-54) | -0,025 | 0,013 | 0,059 | 0,099 |  | -0,049 | 0,024 | 0,041 | 0,074 |  | -0,093 | 0,028 | 0,001 | 0,003 |  | 0,030 | 0,025 | 0,227 | 0,300 |
| 65 or more (vs 45-54) | -0,122 | 0,056 | 0,031 | 0,060 |  | 0,173 | 0,103 | 0,095 | 0,155 |  | -0,280 | 0,123 | 0,022 | 0,045 |  | -0,146 | 0,109 | 0,182 | 0,250 |
| **Education** (vs upper secondary/vocational) |  |  |  |  |  |  |  |  |  |  |  |  |  |  |  |  |  |  |  |
| Compulsory | 0,153 | 0,022 | 0,000 | 0,000 |  | -0,103 | 0,040 | 0,010 | 0,022 |  | 0,068 | 0,047 | 0,149 | 0,215 |  | -0,051 | 0,042 | 0,225 | 0,300 |
| Bachelor’s/university degree | -0,093 | 0,008 | 0,000 | 0,000 |  | 0,089 | 0,014 | 0,000 | 0,000 |  | 0,010 | 0,017 | 0,537 | 0,608 |  | 0,023 | 0,015 | 0,133 | 0,197 |
| Master/PhD | -0,120 | 0,008 | 0,000 | 0,000 |  | 0,129 | 0,015 | 0,000 | 0,000 |  | 0,020 | 0,018 | 0,274 | 0,356 |  | 0,029 | 0,016 | 0,071 | 0,117 |
| **BMI (kg/m^2)** (vs <25 (Underweight or normal)) |  |  |  |  |  |  |  |  |  |  |  |  |  |  |  |  |  |  |  |
| 25-30 (Overweight) | 0,003 | 0,008 | 0,700 | 0,780 |  | -0,002 | 0,014 | 0,890 | 0,948 |  | -0,011 | 0,016 | 0,484 | 0,556 |  | -0,024 | 0,014 | 0,100 | 0,159 |
| >30 (Obese) | 0,034 | 0,010 | 0,001 | 0,003 |  | 0,056 | 0,018 | 0,002 | 0,005 |  | 0,000 | 0,021 | 0,991 | 0,991 |  | -0,088 | 0,019 | 0,000 | 0,000 |
| **History of psychiatric disorders** |  |  |  |  |  |  |  |  |  |  |  |  |  |  |  |  |  |  |  |
| Yes vs No | 0,170 | 0,009 | 0,000 | 0,000 |  | 0,061 | 0,016 | 0,000 | 0,001 |  | 0,039 | 0,019 | 0,045 | 0,078 |  | 0,005 | 0,017 | 0,750 | 0,824 |
| **Living alone** |  |  |  |  |  |  |  |  |  |  |  |  |  |  |  |  |  |  |  |
| Yes vs No | 0,277 | 0,030 | 0,000 | 0,000 |  | 0,136 | 0,055 | 0,013 | 0,027 |  | -0,206 | 0,065 | 0,002 | 0,004 |  | -0,088 | 0,058 | 0,134 | 0,197 |
| **Chronic medical conditions** |  |  |  |  |  |  |  |  |  |  |  |  |  |  |  |  |  |  |  |
| 1 condition (vs no) | 0,158 | 0,008 | 0,000 | 0,000 |  | -0,061 | 0,015 | 0,000 | 0,000 |  | 0,002 | 0,018 | 0,929 | 0,967 |  | -0,057 | 0,016 | 0,000 | 0,001 |
| 2 conditions (vs no) | 0,322 | 0,021 | 0,000 | 0,000 |  | -0,154 | 0,038 | 0,000 | 0,000 |  | -0,009 | 0,045 | 0,841 | 0,911 |  | -0,084 | 0,040 | 0,036 | 0,066 |
| > 2 conditions (vs no) | 0,296 | 0,047 | 0,000 | 0,000 |  | -0,086 | 0,087 | 0,320 | 0,402 |  | 0,073 | 0,103 | 0,476 | 0,556 |  | -0,148 | 0,092 | 0,108 | 0,168 |
| **Income lost due to the pandemic** |  |  |  |  |  |  |  |  |  |  |  |  |  |  |  |  |  |  |  |
| yes, some (vs no) |  |  |  |  |  | 0,029 | 0,014 | 0,035 | 0,065 |  | 0,028 | 0,018 | 0,110 | 0,169 |  | -0,035 | 0,016 | 0,033 | 0,063 |
| yes, significant (vs no) |  |  |  |  |  | 0,116 | 0,020 | 0,000 | 0,000 |  | -0,034 | 0,025 | 0,176 | 0,245 |  | -0,150 | 0,023 | 0,000 | 0,000 |
| **Quarantined/isolated** |  |  |  |  |  |  |  |  |  |  |  |  |  |  |  |  |  |  |  |
| Yes (vs no) |  |  |  |  |  | 0,080 | 0,010 | 0,000 | 0,000 |  | 0,107 | 0,010 | 0,000 | 0,000 |  | 0,001 | 0,017 | 0,961 | 0,978 |
| **SARS-CoV-2-infection** |  |  |  |  |  |  |  |  |  |  |  |  |  |  |  |  |  |  |  |
| Yes (vs no) |  |  |  |  |  | 0,048 | 0,069 | 0,481 | 0,556 |  | -0,006 | 0,049 | 0,899 | 0,948 |  | 0,097 | 0,036 | 0,006 | 0,014 |
| **Vaccination (SARS-CoV-2) status** |  |  |  |  |  |  |  |  |  |  |  |  |  |  |  |  |  |  |  |
| Yes (vs no) |  |  |  |  |  |  |  |  |  |  | -0,184 | 0,030 | 0,000 | 0,000 |  | 0,014 | 0,017 | 0,404 | 0,492 |
| **Prepandemic mental distress** | 0,197 | 0,002 | 0,000 | 0,000 |  | 0,005 | 0,003 | 0,161 | 0,229 |  | 0,043 | 0,004 | 0,000 | 0,000 |  | -0,041 | 0,004 | 0,000 | 0,000 |

Notes: Model was estimated with full Information Maximum Likelihood method, BMI: Body Mass Index. SE: Standard error; FDR p: False Discovery Rate corrected p-value.
